# A Multi-Cohort nnU-Net for White Matter Hyperintensity Segmentation Across 2D and 3D FLAIR Acquisitions

**DOI:** 10.64898/2026.11.11.26367656

**Authors:** Yasheng Chen, Uttam Khatri, Byron Xu, Serguei V. Astafiev, Andria Ford, Jin-Moo Lee, Hongyu An, Chia-Ling Phuah

## Abstract

**Background:** White matter hyperintensities (WMH) are increasingly pooled across aging, cerebral small vessel disease (CSVD), and dementia cohorts that differ in both disease composition and FLAIR acquisition. Most automated segmentation tools were developed within a narrower range of populations or protocols, and few have been tested for whether segmentation accuracy and error remain consistent across conventional 2D and isotropic 3D FLAIR, or whether the choice of algorithm alters downstream clinical associations.

**Methods:** We developed a single nnU-Net model, taking FLAIR and T1-weighted images as inputs, using 460 2D and 3D FLAIR scans from a neurodegenerative cohort (Alzheimer’s Disease Neuroimaging Initiative, ADNI), a CSVD-enriched cohort with same-visit paired 2D and 3D FLAIR (Washington University White Matter Ischemic-Hypoxia study, WWISH), and a population-based sample (UK Biobank), holding out 92 scans from 66 participants at the participant-level as an internal test set. We further evaluated the model in an independent ADNI3 3D FLAIR set (n = 25), benchmarking voxel-wise overlap (Dice), volumetric agreement, and lesion-level detection against SHIVA-WMH, segcsvdWMH, and FreeSurfer WMH-SynthSeg, and compared acquisition-specific volume bias in participants with paired 2D and 3D FLAIR. We then compared how WMH volumes from each method predicted incident dementia in cognitively unimpaired adults from ADNI and the Knight Alzheimer Disease Research Center (n = 1,174; 42 incident cases).

**Results:** In the internal test set, the model achieved a mean Dice of 0.73 (SD 0.10), compared with 0.50, 0.57, and 0.29 for SHIVA-WMH, segcsvdWMH, and WMH-SynthSeg (all FDR-adjusted *p* < 0.001). Dice was similar on 2D and 3D FLAIR (0.75 vs. 0.71). In WWISH participants imaged with both acquisitions, mean volume bias differed between 2D and 3D scans by 1.8 mL for the model versus 4.7–9.9 mL for the comparators. Advantages were largest in low-burden scans (<5 mL; Dice 0.66 vs. 0.19–0.54) and for lesions of ≤100 voxels (detection 77–87% vs. 14–63%). In ADNI3, the model retained a mean Dice of 0.70 (SD 0.14) without retraining and exceeded all comparators (FDR-adjusted *p* ≤ 0.002). Model-derived WMH volume was associated with incident dementia beyond clinical covariates (OR per SD 1.40, 95% CI 1.11–1.77; likelihood-ratio *p* = 0.009; cross-validated AUC 0.71 vs. 0.68), whereas comparator-derived volumes did not significantly improve model fit.

**Conclusions:** Training a single model across disease contexts and FLAIR acquisitions yielded WMH segmentation that was similarly accurate on 2D and 3D FLAIR, sensitive to low-burden disease, transferable to an unseen protocol, and clinically informative, supporting its use in multi-cohort, mixed-etiology studies.

**KEY POINTS:**

- A single nnU-Net trained across neurodegenerative, cerebral small vessel disease, and population-based cohorts and across 2D and 3D FLAIR outperformed SHIVA-WMH, segcsvdWMH, and WMH-SynthSeg, with the largest gains in low-burden scans and small lesions.
- In same-visit paired 2D and 3D FLAIR scans, the model’s volume bias relative to acquisition-specific reference masks varied less across acquisitions, whereas comparator biases varied with acquisition.
- Only WMH volume from the model significantly improved model fit for incident dementia beyond clinical covariates, highlighting the potential influence of segmentation algorithm choice on downstream clinical inference.

## INTRODUCTION

White matter hyperintensities (WMH) are among the most common neuroimaging findings in older adults and are associated with cerebral small vessel disease (CSVD), cognitive decline, stroke, and dementia. Although WMH are often interpreted as markers of vascular brain injury, their biological substrates are heterogeneous and may reflect contributions from small vessel injury, neurodegeneration, and other vascular and metabolic processes (Wardlaw et al., 2015; Smith and Beaudin, 2018; Vemuri et al., 2022; Maillard et al., 2024). Accordingly, WMH are increasingly quantified as a continuous marker of cumulative white matter injury across population-based aging studies, vascular cohorts, and studies of Alzheimer’s disease and related dementias (AD/ADRD) (Garnier-Crussard et al., 2023; DeCarli et al., 2025). Reliable automated segmentation is essential for these applications because manual delineation is labor-intensive, subject to inter-rater variability, and impractical for large, multisite imaging datasets.

The broader use of WMH as a cross-disease imaging biomarker, however, presents a substantial translational challenge because models are often applied to populations that differ from their development cohorts in both clinical characteristics and MRI acquisition protocols. Contemporary neuroimaging studies frequently combine participants with different disease profiles and lesion burdens whose scans were acquired on different scanners and with different FLAIR protocols, including conventional thick-slice 2D and approximately isotropic 3D sequences. Segmentation performance can vary across these conditions, particularly in participants with low WMH burden or small punctate lesions, and protocol-dependent errors may introduce systematic differences in estimated WMH volume (Heinen et al., 2019; Hotz et al., 2022; Kuijf et al., 2019; Tubi et al., 2020; Torres-Simon et al., 2024). Methods developed within a particular disease spectrum or acquisition setting may therefore perform less consistently when both the population and imaging protocol differ from their development distributions (Caligiuri et al., 2015; Balakrishnan et al., 2021; Rahmani et al., 2024). This mismatch complicates pooled analyses and may force investigators to use different algorithms across sub-cohorts, exclude incompatible acquisitions, or deploy a single method beyond the setting in which it was validated.

Recent deep-learning methods have substantially advanced automated WMH segmentation. SHIVA-WMH has demonstrated sensitivity to early WMH in vascular and aging cohorts, segcsvdWMH was developed using heterogeneous datasets to quantify WMH in CSVD and related clinical populations, and WMH-SynthSeg was designed to support segmentation across heterogeneous clinical MRI data (Gibson et al., 2024; Laso et al., 2024; Tsuchida et al., 2024). These methods address important dimensions of population or acquisition heterogeneity. However, evidence remains limited for a single empirically trained model designed and evaluated across the intersection of neurodegenerative, vascular, and population-based cohorts and both conventional 2D and isotropic 3D FLAIR acquisitions. Moreover, relatively few studies have determined whether performance is maintained across lesion burden and lesion size, tested transfer to an unseen acquisition configuration, or examined whether algorithm-dependent differences in WMH quantification alter downstream clinical information (Tubi et al., 2020; Gesierich et al., 2025). Addressing this intersectional gap is necessary to establish a consistent WMH measure for large, heterogeneous neuroimaging studies.

In this study, we used the self-configuring nnU-Net framework (Isensee et al., 2021) to develop a single WMH segmentation model using three complementary cohorts: the Alzheimer’s Disease Neuroimaging Initiative (ADNI), representing the neurodegenerative disease spectrum; the Washington University White Matter Ischemic-Hypoxia study (WWISH), enriched for CSVD and contributing paired 2D and 3D FLAIR acquisitions; and UK Biobank (UKB), representing population-based aging. Together, these cohorts encompass substantial variation in WMH burden, lesion morphology, disease context, and FLAIR acquisitions. We addressed four questions: 1) whether the model provides accurate and consistent overall segmentation across cohorts and acquisition types relative to contemporary benchmark methods; 2) whether it maintains sensitivity to small lesions and low WMH burden; 3) whether the model maintains its performance on an independent set of ADNI 3D FLAIR scans not used for model development or tuning; and 4) whether its WMH volume estimates provide incremental information about incident dementia beyond established clinical predictors. We hypothesized that deliberately incorporating heterogeneity in disease context, lesion burden, and acquisition into model development would produce a single deployable model with consistent segmentation performance across the evaluated imaging settings and clinically informative WMH estimates.

## METHODS

### Participants and imaging datasets

We assembled a multi-cohort imaging dataset designed to reflect the heterogeneity of populations and FLAIR acquisitions encountered in aging and cerebrovascular research. Participants were drawn from three cohorts: the Alzheimer’s Disease Neuroimaging Initiative (ADNI), representing an AD-focused neurodegenerative cohort; the Washington University White Matter Ischemic-Hypoxia study (WWISH), representing a CSVD-enriched cohort; and the UK Biobank (UKB), representing population-based aging. Together, these cohorts encompassed a broad range of WMH burden, lesion morphology, clinical context, and 2D and 3D FLAIR acquisition protocols.

The model-development dataset comprised 460 FLAIR scans from 330 unique participants. ADNI contributed one 2D FLAIR scan from each of 100 participants spanning cognitively unimpaired, mild cognitive impairment, and dementia groups. WWISH contributed same-visit paired 2D and 3D FLAIR scans from 130 participants with clinically defined CSVD, for a total of 260 scans. UKB contributed one 3D FLAIR scan from each of 100 community-dwelling participants, generally characterized by lower WMH burden.

The dataset was partitioned using participant-level grouping, such that each participant was assigned exclusively to the training, validation, or internal test set. For WWISH participants, paired 2D and 3D FLAIR scans were assigned together to the same partition, preventing subject-level information leakage across partitions. The internal held-out test set comprised 92 scans from 66 unique participants: 20 ADNI participants with one 2D FLAIR scan each, 26 WWISH participants with paired 2D and 3D FLAIR scans, and 20 UKB participants with one 3D FLAIR scan each. The remaining 368 scans from 264 participants formed the model-development set used for training and validation. Within the development set, participant-level grouping was maintained throughout model fitting and validation so that both WWISH acquisitions from a given participant remained in the same subset (training or validation). External validation was performed in an independent ADNI3 test cohort of 25 nonoverlapping participants with 3D FLAIR imaging that was entirely withheld from model development and tuning.

Demographic and clinical characteristics of ADNI and WWISH participants and the distributions of WMH volume across all three development cohorts are summarized in Table 1. Linked demographic and clinical data were not available for this UKB subset. Cohort-specific inclusion criteria are described in the **Supplementary Methods**, and FLAIR acquisition parameters are provided in **Supplementary Table S1**.

**Table 1.**
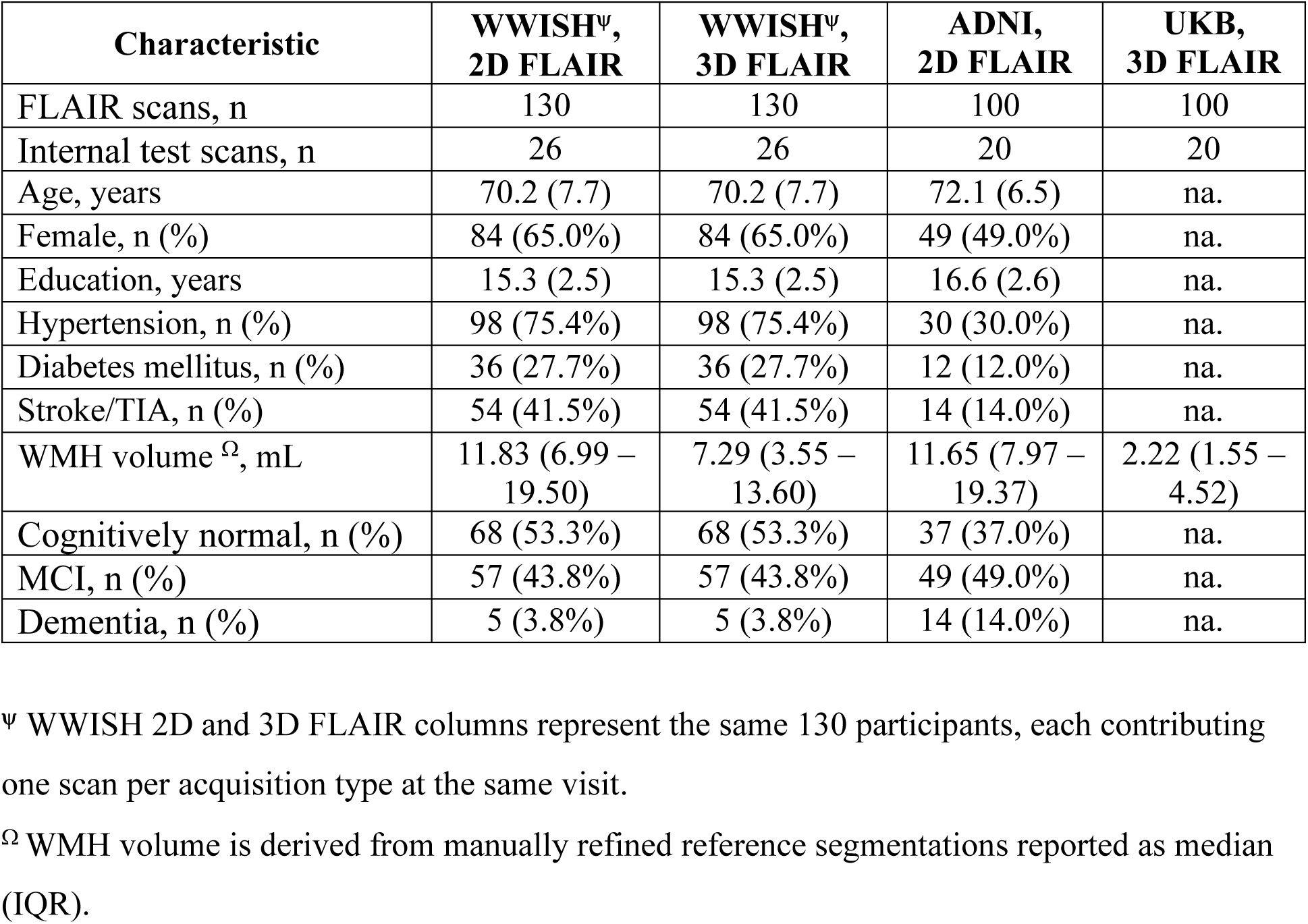
Cohort characteristics for WMH segmentation model development and internal testing. Continuous variables are reported as mean (SD), categorical variables as n (%). Demographic and clinical data were unavailable for the UKB subset; Cognitive status was based on the CDR for ADNI and the MoCA for WWISH. ADNI, Alzheimer’s Disease Neuroimaging Initiative; CDR, Clinical Dementia Rating; MCI, mild cognitive impairment; MoCA, Montreal Cognitive Assessment; TIA, transient ischemic attack; UKB, UK Biobank; WMH, white matter hyperintensity; WWISH, Washington University White Matter Ischemic-Hypoxia study.

| Characteristic | **WWISH<sup>Ψ</sup>, 2D FLAIR** | **WWISH<sup>Ψ</sup>, 3D FLAIR** | **ADNI, 2D FLAIR** | **UKB, 3D FLAIR** |
|---|---|---|---|---|
| FLAIR scans, n | 130 | 130 | 100 | 100 |
| Internal test scans, n | 26 | 26 | 20 | 20 |
| Age, years | 70.2 (7.7) | 70.2 (7.7) | 72.1 (6.5) | na. |
| Female, n (%) | 84 (65.0%) | 84 (65.0%) | 49 (49.0%) | na. |
| Education, years | 15.3 (2.5) | 15.3 (2.5) | 16.6 (2.6) | na. |
| Hypertension, n (%) | 98 (75.4%) | 98 (75.4%) | 30 (30.0%) | na. |
| Diabetes mellitus, n (%) | 36 (27.7%) | 36 (27.7%) | 12 (12.0%) | na. |
| Stroke/TIA, n (%) | 54 (41.5%) | 54 (41.5%) | 14 (14.0%) | na. |
| WMH volume<sup>Ω</sup>, mL | 11.83 (6.99 – 19.50) | 7.29 (3.55 – 13.60) | 11.65 (7.97 – 19.37) | 2.22 (1.55 – 4.52) |
| Cognitively normal, n (%) | 68 (53.3%) | 68 (53.3%) | 37 (37.0%) | na. |
| MCI, n (%) | 57 (43.8%) | 57 (43.8%) | 49 (49.0%) | na. |
| Dementia, n (%) | 5 (3.8%) | 5 (3.8%) | 14 (14.0%) | na. |

<sup>Ψ</sup> WWISH 2D and 3D FLAIR columns represent the same 130 participants, each contributing one scan per acquisition type at the same visit.
<sup>Ω</sup> WMH volume is derived from manually refined reference segmentations reported as median (IQR).

### WMH reference segmentation

Reference WMH masks were generated using a standardized semiautomated labeling strategy. An existing automated WMH segmentation method (Phuah et al., 2022) was first applied to each FLAIR image to provide an initial candidate mask. A trained rater then reviewed and manually refined each mask using threshold-guided and voxel-wise editing to correct false-positive regions, recover missed lesions, and improve lesion-boundary delineation. Final masks underwent review and adjudication by an experienced neurologist. For WWISH participants, reference masks were generated separately for the 2D and 3D FLAIR acquisitions. Consequently, each acquisition was evaluated against its acquisition-specific reference segmentation.

### Image preprocessing

All images underwent a standardized preprocessing pipeline to reduce nonbiological variability across cohorts and acquisition types, adapted from a previously described approach (Phuah et al., 2022). T1-weighted images were first resampled to 1-mm isotropic resolution. T1-weighted and FLAIR images were then skull-stripped using FreeSurfer SynthStrip (Hoopes et al., 2022) and corrected for intensity inhomogeneity using N4-bias field correction in ANTs (Tustison et al., 2010). Both 2D and 3D FLAIR images were registered to their corresponding resampled T1-weighted images using affine registration in FSL-FLIRT (Jenkinson et al., 2002). The resulting transformation was applied to resample each FLAIR image and its WMH reference mask onto the 1-mm isotropic T1-weighted image grid. Within the T1-derived SynthStrip brain mask, T1-weighted and registered FLAIR intensities were normalized by subtracting the mean and dividing by three times the standard deviation, then clipped to [-1,1]. The same workflow was applied across cohorts and acquisition types. **Figure 1** summarizes the preprocessing, model development and evaluation workflow.

**Figure 1.**
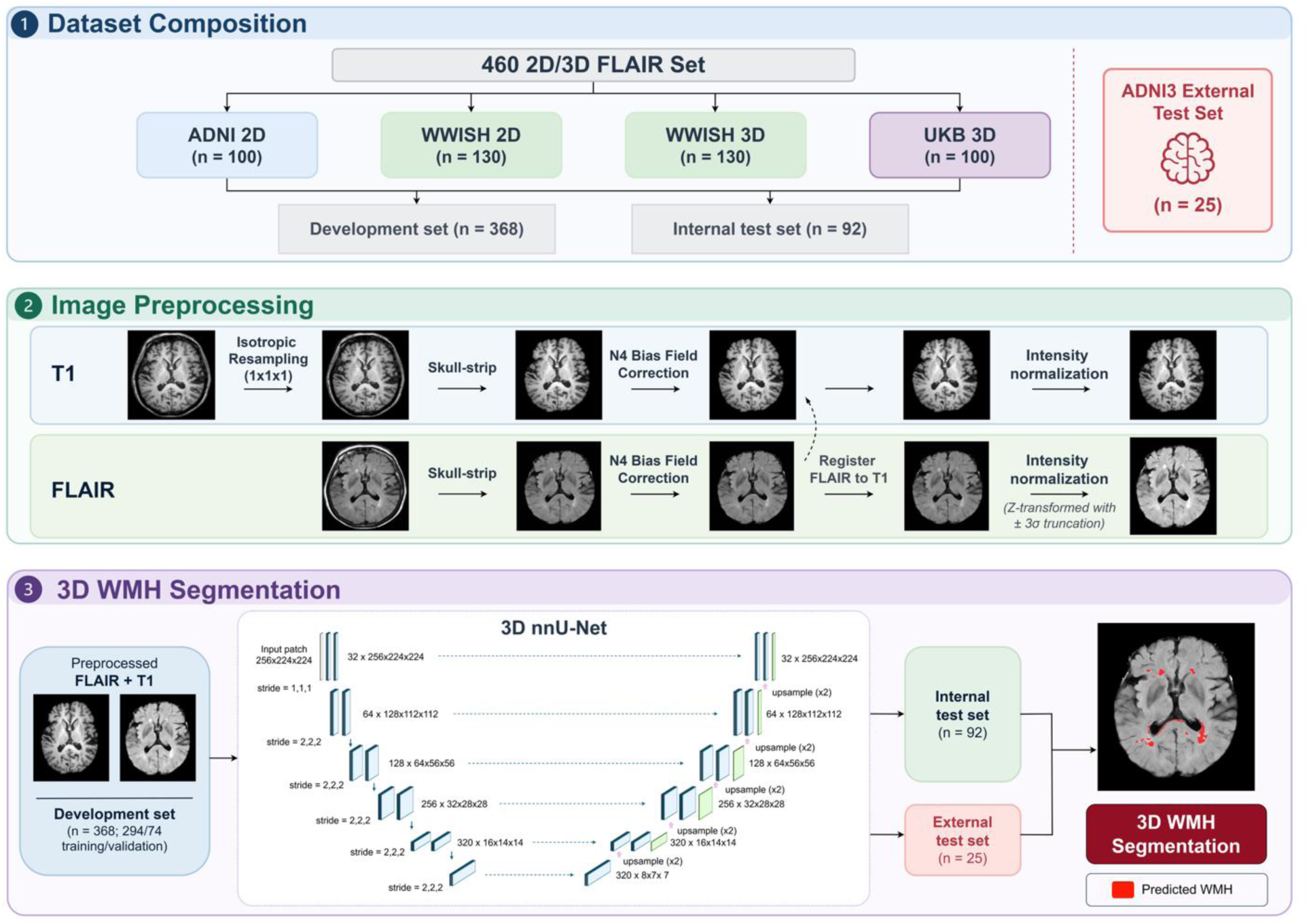
Study design and processing workflow. ***Top:* Dataset composition and partitioning.** The model-development dataset comprised 460 FLAIR scans from 330 participants: 100 ADNI 2D scans, paired same-visit 2D and 3D scans from 130 WWISH participants (260 scans), and 100 UKB 3D scans. Participant-level splitting yielded 368 development scans (294 training, 74 validation) and 92 internal test scans, with paired WWISH acquisitions retained in the same subset. An independent ADNI3 set of 25 3D scans was reserved for external testing. ***Middle:* Image preprocessing.** T1-weighted and FLAIR images underwent skull stripping, N4 bias-field correction, affine coregistration, resampling to a 1-mm isotropic T1-weighted grid, and within-brain intensity normalization using a common pipeline across cohorts and acquisition types. ***Bottom:* WMH segmentation.** A single 3D nnU-Net with FLAIR and T1-weighted input channels was applied to both test sets without retraining. Network labels indicate feature channels × feature-map dimensions; strides indicate downsampling factors. Predicted WMH are shown in red. Abbreviations: ADNI, Alzheimer’s Disease Neuroimaging Initiative; FLAIR, fluid-attenuated inversion recovery; UKB, UK Biobank; WMH, white matter hyperintensity; WWISH, Washington University White Matter Ischemic-Hypoxia study.

### Model architecture and training

We implemented a single 3D nnU-Net model for WMH segmentation (Isensee et al., 2021). The network received two input channels: the preprocessed FLAIR image and the coregistered T1-weighted image. nnU-Net automatically configures network architecture, patch size, batch size, and other training parameters according to the properties of the input dataset. The resulting model used an encoder-decoder architecture with skip connections, instance normalization, and leaky rectified linear unit activations.

Both native 2D and native 3D FLAIR acquisitions were represented as 3D image volumes after preprocessing and resampling, and coregistration to the T1-weighted image. The same network was therefore applied to both acquisition types; separate 2D- and 3D-specific networks were not trained. This design was intended to produce a single model deployable across the range of FLAIR protocols represented in the development data.

A central feature of the training strategy was the deliberate integration of three complementary cohorts spanning neurodegenerative disease, CSVD, and population-based aging, together with both 2D and 3D FLAIR acquisitions. This design exposed a single model to variation in clinical context, WMH burden and morphology, and FLAIR acquisition during training, rather than developing separate disease- or sequence-specific models. Training patches were sampled from the preprocessed FLAIR and T1-weighted images and corresponding reference masks and underwent the standard nnU-Net on-the-fly spatial and intensity augmentations. The model was optimized using a composite Dice and cross-entropy loss to address the substantial class imbalance between WMH and non-WMH voxels.

The model development set was randomly divided at the participant level into a training set (294 scans from 211 participants) and a validation set (74 scans from 53 participants), with paired WWISH 2D and 3D scans from the same participant assigned to the same subset. A single model was trained on the training set, and the validation set was used to monitor training and select the final checkpoint. At inference, the model’s *softmax* outputs was binarized using a probability threshold of 0.5. The internal test set and ADNI3 3D set remained excluded from all model fitting and model-selection procedures.

### Benchmark algorithms

The proposed model was benchmarked against three contemporary automated WMH segmentation methods: SHIVA-WMH (Tsuchida et al., 2024), segcsvdWMH (Gibson et al., 2024), and FreeSurfer WMH-SynthSeg (Laso et al., 2024). These methods were selected to represent recently developed approaches intended for vascular, heterogeneous clinical, or large-scale imaging applications. Each benchmark method was run using its recommended configuration and decision threshold: 0.5 for SHIVA-WMH, 0.35 for segcsvdWMH, and the discrete WMH label provided by WMH-SynthSeg. Each method received its recommended inputs: FLAIR and T1-weighted for SHIVA-WMH, and FLAIR for both segcsvdWMH and WMH-SynthSeg. Input preprocessing required by each method was applied accordingly. Algorithm outputs were subsequently resampled and aligned to the corresponding reference-mask space to permit evaluation using common voxel- and lesion-level definitions. All methods were evaluated on the same internal test and ADNI3 validation acquisitions. No benchmark method was retrained or fine-tuned using the internal test or ADNI3 data.

### Segmentation performance

Segmentation performance was evaluated at the voxel, volume, and lesion levels. The Dice similarity coefficient between the predicted and reference WMH masks was the primary voxel-level metric. Volumetric performance was evaluated using signed and absolute differences between predicted and reference total WMH volumes. Agreement across the range of WMH burden was further examined using Bland-Altman analysis. For lesion-level evaluation, spatially connected WMH components were identified separately in the predicted and reference masks. A reference lesion was considered detected if it shared at least one voxel with a predicted lesion (Ganiler et al., 2014). For each scan, we calculated the proportion of reference lesions detected, lesion-level Dice, and the number of false-positive predicted lesions. Lesions were stratified by reference volume into four prespecified categories: very small, 15–30 voxels; small, 31–100 voxels; medium, 101–250 voxels; and large, > 250 voxels. Because all masks were represented at 1-mm isotropic resolution, one voxel corresponded to 1mm^3^.

Performance was examined overall and within prespecified strata defined by cohort, native FLAIR acquisition type, reference WMH burden, and lesion size. WMH burden was categorized as low, < 5 mL; moderate, 5–10 mL; or high, > 10 mL. Acquisition-specific analyses included all 2D scans, all 3D scans, and cohort-by-acquisition subgroups comprising ADNI 2D, WWISH 2D, WWISH 3D, UKB 3D, and external ADNI3 3D. As secondary analysis, we assessed direct agreement between predicted WMH volumes from same-visit paired 2D and 3D FLAIR scans in 26 WWISH participants from the held-out internal test set. The same analyses were applied to the paired expert reference volumes (**Supplementary Table S5**).

### Cognitive outcome analysis

We evaluated the downstream clinical validity of WMH volume estimates by examining their associations with incident dementia among cognitively unimpaired participants (global Clinical Dementia Rating [CDR] score = 0; Morris et al., 1997) from ADNI and the Knight Alzheimer Disease Research Center (Knight ADRC) cohort. Participants were eligible if they had a baseline FLAIR scan and a CDR assessment within 180 days of imaging. Incident dementia was defined as progression to a global CDR of ≥1 during follow-up. Both cohorts included baseline 2D and 3D FLAIR scans; ADNI training participants were excluded to prevent participant-level overlap.

For each segmentation method, total WMH volume was calculated as the number of WMH-labelled voxels multiplied by voxel volume. Separate logistic regression models evaluated whether WMH volume derived from each algorithm provided incremental information about incident dementia beyond age, sex, years of education, *APOE ε*4 carrier status, and cohort. A clinical-only model containing these covariates served as the reference; each algorithm-specific model additionally included standardized WMH volume. Associations were summarized as odds ratios per 1-SD increase in WMH volume, with 95% confidence intervals, and Wald *p*-values. Incremental model fit was assessed using likelihood-ratio tests comparing each WMH-augmented model with the clinical-only model. Because the sample included both 2D and 3D FLAIR, and methods differed in acquisition-dependent volume offsets, a sensitivity analysis additionally adjusted both the clinical and WMH-augmented models for FLAIR acquisition type.

Discrimination was assessed using 10-fold cross-validated area under the receiver operating characteristic (AUC) curve. The same 10-fold splits were used for all algorithm-specific models, ensuring that cross-validated AUCs were evaluated in identical held-out participants. Confidence intervals were estimated by bootstrap resampling. Permutation testing with randomly permuted outcome labels was used to assess the significance of ΔAUC relative to the clinical model.

### Statistical analysis

Segmentation metrics were summarized at the scan level. Differences among segmentation methods were evaluated using the Friedman test, followed by paired Wilcoxon signed-rank tests comparing the proposed model with each benchmark method. These analyses were conducted in the overall internal test set and within prespecified acquisition, cohort-by-acquisition, WMH burden, and lesion size strata. For each metric, Wilcoxon *p*-values were adjusted jointly across all strata and all three comparator comparisons using the Benjamini-Hochberg false-discovery-rate (FDR) procedure. The overall internal test set included paired 2D and 3D acquisitions from 26 WWISH participants and therefore contained repeated observations from these participants. Performance was consequently also evaluated separately by acquisition and cohort-by-acquisition stratum, within which each participant contributed one scan. The pooled scan-level analyses did not explicitly account for within-participant correlation between paired WWISH acquisitions; the acquisition-specific analyses were therefore used to assess the robustness of the pooled findings. Cross-acquisition volumetric agreement was assessed using absolute-agreement intraclass correlation coefficients (ICCs) with 95% confidence intervals and Bland-Altman analysis. Paired differences were defined as predicted 3D minus 2D WMH volume; mean bias and 95% limits of agreement (mean bias ±1.96 SD of the paired differences) were reported. All analyses were performed in Python version 3.13.5 using NumPy version 2.1.3; Pandas version 2.2.3; SciPy version 1.15.3; statsmodels version 0.14.4.

## RESULTS

### Cohort and acquisition characteristics

By design, the development cohorts differed in WMH burden, vascular risk profile, and FLAIR acquisition (**Table 1**). Mean reference WMH volume was lowest in UKB (3.6 mL; 3D FLAIR only) and higher in ADNI (15.5 mL; 2D FLAIR only) and WWISH, which also had the highest prevalence of hypertension (75.4%), diabetes (27.7%), and prior stroke or TIA (41.5%). In WWISH, the same 130 participants had a mean reference volume of 16.1 mL on 2D FLAIR and 11.2 mL on 3D FLAIR, illustrating that measured WMH volume depends on acquisition even when masks are expert-refined. Characteristics of the incident dementia analysis sample are summarized in **Table 3**.

### Internal performance and benchmarking

In the internal held-out test set (92 scans from 66 participants), the proposed model achieved a mean Dice of 0.73 (SD 0.10; median 0.72, IQR 0.67–0.81). Mean Dice was 0.50 for SHIVA-WMH, 0.57 for segcsvdWMH, and 0.29 for WMH-SynthSeg (Friedman *p* < 0.001; FDR-adjusted Wilcoxon *p* ≤ 5.3 × 10^−16^ for each comparison with the proposed model; **Table 2**). The proposed model also had the smallest volumetric error (median absolute difference 0.96 mL [IQR 0.36–2.12] vs. 2.17–3.36 mL for comparators) and the smallest mean bias (−0.86 mL [SD 3.11] vs. −1.83 to −3.90 mL; **Supplementary Table S2**). Bland-Altman analysis showed narrower limits of agreement for the proposed model than for the comparators, with modest underestimation in a small number of high-burden 2D scans (**Supplementary Figure S1**).

**Table 2.** Voxel-wise Dice and absolute WMH volume error by segmentation method in the internal held-out test set and the external ADNI3 validation set. Dice values are mean (95% CI) and absolute volume errors as median (IQR), in mL. Absolute volume error was calculated as the absolute difference between predicted and reference total WMH volumes. Adjusted *p* values are from paired Wilcoxon signed-rank tests comparing each method with the proposed nnU-Net model, with Benjamini–Hochberg FDR correction. The all-scan internal analysis include paired 2D and 3D acquisitions from 26 WWISH participants; within cohort-by-acquisition strata, each participant contributes one scan. The ADNI3 set was not used for training, model selection, or threshold tuning. ADNI, Alzheimer’s Disease Neuroimaging Initiative; CI, confidence interval; FDR, false discovery rate; FLAIR, fluid-attenuated inversion recovery; UKB, UK Biobank; WWISH, Washington University White Matter Ischemic-Hypoxia study.

| **Analysis group** | **N scans** | **Method** | **Dice,<br>mean (95% CI)** | **Adjusted *P* value** | **Absolute volume error, mL, median (IQR)** |
|---|---|---|---|---|---|
| **All<br>(2D + 3D FLAIR)** | 92 | *nnUNet* | 0.73 (0.71, 0.75) | na | 0.96 (0.36 – 2.12) |
|  | 92 | *SHIVA* | 0.50 (0.46, 0.54) | 1.63 x 10<sup>-16</sup> | 2.25 (1.12 – 5.07) |
|  | 92 | *segCSVD* | 0.57 (0.54, 0.61) | 5.29 x 10<sup>-16</sup> | 2.17 (0.76 – 5.54) |
|  | 92 | *SynthSeg* | 0.29 (0.26, 0.33) | 1.63 x 10<sup>-16</sup> | 3.36 (1.39 – 6.68) |
| **2D FLAIR only** | 46 | *nnUNet* | 0.75 (0.72, 0.77) | na | 1.59 (0.88 – 3.39) |
|  | 46 | *SHIVA* | 0.52 (0.46, 0.57) | 4.26 x 10<sup>-14</sup> | 3.70 (2.62 – 5.91) |
|  | 46 | *segCSVD* | 0.54 (0.48, 0.59) | 4.26 x 10<sup>-14</sup> | 5.31 (3.56 – 8.88) |
|  | 46 | *SynthSeg* | 0.30 (0.25, 0.35) | 4.26 x 10<sup>-14</sup> | 5.16 (2.30 – 10.42) |
| **3D FLAIR only** | 46 | *nnUNet* | 0.71 (0.67, 0.74) | na | 0.55 (0.19 – 1.03) |
|  | 46 | *SHIVA* | 0.48 (0.41, 0.55) | 4.23 x 10<sup>-9</sup> | 1.21 (0.66 – 1.97) |
|  | 46 | *segCSVD* | 0.61 (0.56, 0.66) | 3.26 x 10<sup>-10</sup> | 0.81 (0.45 – 1.26) |
|  | 46 | *SynthSeg* | 0.29 (0.23, 0.35) | 4.23 x 10<sup>-9</sup> | 2.31 (1.16 – 3.74) |
| **ADNI<br>(2D FLAIR)** | 20 | *nnUNet* | 0.73 (0.69, 0.78) | na | 2.34 (0.86 – 5.11) |
|  | 20 | *SHIVA* | 0.46 (0.38, 0.55) | 2.29 x 10<sup>-6</sup> | 3.85 (2.75 – 10.67) |
|  | 20 | *segCSVD* | 0.46 (0.38, 0.54) | 2.29 x 10<sup>-6</sup> | 6.91 (3.72 – 12.76) |
|  | 20 | *SynthSeg* | 0.19 (0.11, 0.27) | 2.29 x 10<sup>-6</sup> | 4.59 (1.19 – 9.37) |
| **WWISH<br>(2D FLAIR)** | 26 | *nnUNet* | 0.76 (0.72, 0.79) | na | 1.27 (0.88 – 2.67) |
|  | 26 | *SHIVA* | 0.56 (0.48, 0.64) | 4.47 x 10<sup>-8</sup> | 3.64 (2.62 – 5.18) |
|  | 26 | *segCSVD* | 0.59 (0.53, 0.66) | 4.47 x 10<sup>-8</sup> | 5.04 (3.56 – 7.16) |
|  | 26 | *SynthSeg* | 0.38 (0.33, 0.43) | 4.47 x 10<sup>-8</sup> | 5.67 (3.67 – 12.46) |
| **WWISH<br>(3D FLAIR)** | 26 | *nnUNet* | 0.74 (0.69, 0.78) | na | 0.89 (0.25 – 1.42) |
|  | 26 | *SHIVA* | 0.55 (0.46, 0.64) | 9.90 x 10<sup>-6</sup> | 1.70 (0.82 – 3.03) |
|  | 26 | *segCSVD* | 0.66 (0.61, 0.72) | 2.69 x 10<sup>-5</sup> | 0.96 (0.58 – 1.67) |
|  | 26 | *SynthSeg* | 0.41 (0.33, 0.48) | 9.95 x 10<sup>-6</sup> | 2.62 (1.34 – 4.22) |
| **UKB<br>(3D FLAIR)** | 20 | *nnUNet* | 0.67 (0.63, 0.71) | na | 0.35 (0.10 – 0.58) |
|  | 20 | *SHIVA* | 0.39 (0.29, 0.49) | 2.86 x 10<sup>-6</sup> | 0.96 (0.62 – 1.23) |
|  | 20 | *segCSVD* | 0.54 (0.47, 0.62) | 2.86 x 10<sup>-6</sup> | 0.62 (0.36 – 1.05) |
|  | 20 | *SynthSeg* | 0.14 (0.09, 0.19) | 2.86 x 10<sup>-6</sup> | 1.78 (0.87 – 2.46) |
| **External Validation Set** |  |  |  |  |  |
| **ADNI3<br>(3D FLAIR)** | 25 | *nnUNet* | 0.70 (0.65, 0.76) | na | 1.59 (0.60 – 3.28) |
|  | 25 | *SHIVA* | 0.60 (0.54, 0.66) | 3.20 x 10<sup>-5</sup> | 3.15 (1.21 – 7.84) |
|  | 25 | *segCSVD* | 0.68 (0.62, 0.73) | 1.63 x 10<sup>-3</sup> | 1.83 (0.45 – 2.80) |
|  | 25 | *SynthSeg* | 0.42 (0.36, 0.48) | 2.68 x 10<sup>-7</sup> | 2.21 (0.74 – 4.40) |

### Segmentation performance across FLAIR acquisitions and cohorts

The proposed model achieved similar segmentation accuracy across acquisition types, with mean Dice of 0.75 (SD 0.09) on 2D FLAIR and 0.71 (SD 0.12) on 3D FLAIR (**Table 2**; **Figure 2D**). The paired WWISH acquisitions provided a within-participant assessment of acquisition sensitivity. In the same 26 participants, the proposed model had a mean Dice of 0.76 on 2D and 0.74 on 3D FLAIR, with mean volume bias relative to acquisition-specific reference masks was −1.64 mL and +0.13 mL, respectively (**Supplementary Table S2**). Comparator biases varied more between acquisitions. segcsvdWMH underestimated volume more on 2D than on 3D FLAIR (−5.73 vs. −1.06 mL); WMH-SynthSeg substantially underestimated volume on 2D but showed near-zero bias on 3D FLAIR (−9.81 vs. +0.11 mL); and SHIVA-WMH shifted from underestimation on 2D to overestimation on 3D FLAIR (−3.58 vs. +2.46 mL). Thus, the between-acquisition difference in mean bias was 1.77 mL for the proposed model, compared with 4.67–9.92 mL for the comparators. A similar pattern was observed across the full internal test set (**Supplementary Table S2**).

**Figure 2.**
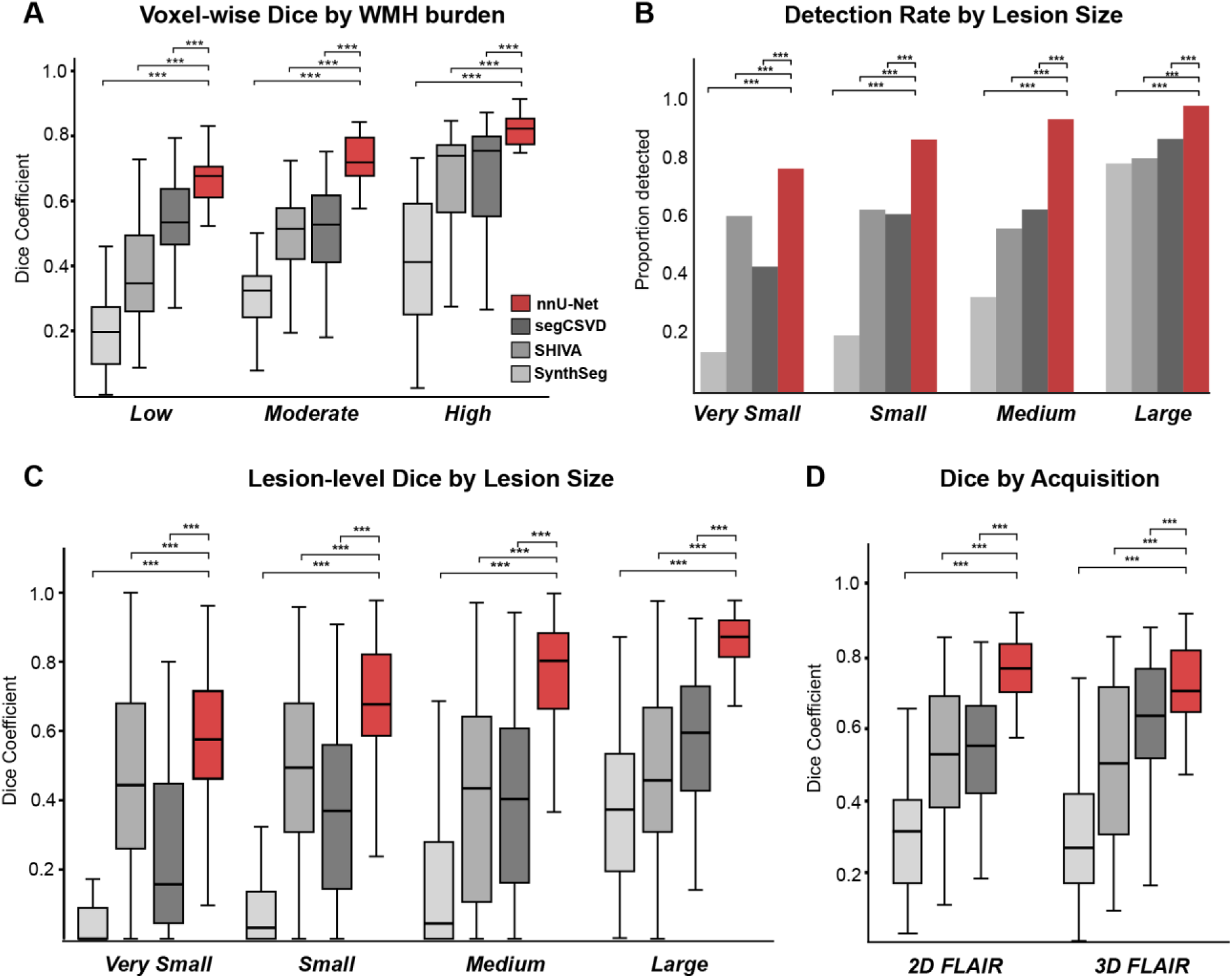
Segmentation performance in the internal held-out test set (92 scans; 66 participants). **A.** Voxel-wise Dice by reference WMH burden (<5, 5–10, and >10 mL). **B.** Lesion detection rate and **C.** lesion-wise Dice by reference lesion size (15–30, 31–100, 101–250, and >250 voxels; 1 voxel = 1 mm^3^). Detection required ≥1 overlapping voxel. **D.** Voxel-wise Dice by native FLAIR acquisition (46 scans each for 2D and 3D). Red, nnU-Net; light-to-dark gray, WMH-SynthSeg, SHIVA-WMH, and segcsvdWMH. Boxes show median and IQR; whiskers extend to 1.5 x IQR. Brackets indicate FDR-adjusted paired Wilcoxon comparisons with nnU-Net (∗ *p* < 0.05, ∗∗ *p* < 0.01, ∗∗∗ *p* < 0.001). Abbreviations: FDR, false discovery rate; FLAIR, fluid-attenuated inversion recovery; WMH, white matter hyperintensity; IQR, interquartile range.

In a secondary analysis, predicted WMH volumes were compared directly between paired 2D and 3D FLAIR scans, independently of the reference masks. The proposed model yielded lower volumes on 3D than on 2D FLAIR (mean difference, −4.57 mL), whereas SHIVA-WMH showed a smaller mean difference (−0.30 mL). Complete agreement statistics, including a separate analysis of paired expert reference volumes, are reported in **Supplementary Table S5**.

Across cohorts, mean Dice for the proposed model was 0.73 in ADNI 2D, 0.76 in WWISH 2D, 0.74 in WWISH 3D, and 0.67 in UKB 3D, and it exceeded each comparator in every cohort-by-acquisition stratum (FDR-adjusted *p* ≤ 2.7 × 10^−5^; **Table 2**; **Supplementary Figure S2**). The lower Dice in UKB is consistent with its low lesion burden, because small absolute errors weigh more heavily on Dice when total WMH volume is small; volumetric error in UKB was the smallest of any cohort (median absolute difference 0.35 mL; **Supplementary Table S2**).

### Performance in low-burden scans and small lesions

Advantages were largest at the low end of the WMH spectrum. Across fixed reference-volume categories, the proposed model’s mean Dice was 0.66 in low-burden (<5 mL), 0.73 in moderate-burden (5–10 mL), and 0.80 in high-burden (>10 mL) scans (**Figure 2A**; **Supplementary Table S3**). In low-burden scans, comparator Dice ranged from 0.19 (WMH-SynthSeg) to 0.54 (segcsvdWMH), and the proposed model exceeded each comparator in all three burden categories (FDR-adjusted *p* < 0.001).

At the lesion level, the proposed model detected 77% of very small (15–30 voxels) and 87% of small (31–100 voxels) reference lesions, compared with 61–63% for SHIVA-WMH, 44–62% for segcsvdWMH, and 14–20% for WMH-SynthSeg (**Figure 2B**; **Supplementary Table S4**). Lesion-wise Dice was also higher in every size category (FDR-adjusted *p* < 0.001; Figure 2C). Higher sensitivity was not achieved by over-segmentation. The proposed model produced fewer false-positive lesions per scan than SHIVA-WMH in every size category (e.g., 0.71 vs. 1.49 very small lesions). segcsvdWMH produced slightly fewer very small false-positive lesions (0.32 per scan), but missed 2.7 times as many very small reference lesions (7.07 vs. 2.62 false negatives per scan). Gains extended to medium lesions (101–250 voxels), for which detection was 94% for the proposed model versus 33–63% for comparators, and to large lesions (>250 voxels; 99% vs. 79–88%).

The small-lesion advantage was largest on 2D FLAIR (**Supplementary Figure S2B**). On 3D FLAIR, SHIVA-WMH, which was developed with isotropic 3D FLAIR to detect punctate lesions, achieved very-small-lesion Dice comparable to that of the proposed model (0.58 vs. 0.64; FDR-adjusted p = 0.66), but with approximately seven times as many very small false-positive lesions per scan (1.98 vs. 0.28; **Supplementary Table S4**).

### External validation in ADNI3

In the independent ADNI3 3D FLAIR set (n = 25), the proposed model achieved a mean Dice of 0.70 (SD 0.14; median 0.75, IQR 0.58–0.80) without retraining or threshold adjustment, comparable to its internal 3D FLAIR performance (**Table 2**; **Figure 3A**). Mean Dice was 0.60 for SHIVA-WMH, 0.68 for segcsvdWMH, and 0.42 for WMH-SynthSeg, and the proposed model exceeded each comparator (FDR-adjusted *p* = 3.2 × 10^−5^, 1.6 × 10^−3^, and 2.7 × 10^−7^, respectively). segcsvdWMH was the closest competitor in this set (median absolute difference 1.83 vs. 1.59 mL for the proposed model; **Supplementary Table S2**), whereas the proposed model modestly overestimated volume (mean bias +1.49 mL). In low-burden ADNI3 scans (<5 mL; n = 10), Dice was similar for the proposed model and segcsvdWMH (0.56 vs. 0.57; FDR-adjusted *p* = 0.49) and higher than for WMH-SynthSeg (0.32; *p* = 0.009; **Supplementary Table S3**). The proposed model detected 86% of very small and small lesions and 99–100% of medium and large lesions (**Figure 3B**; **Supplementary Table S4**). Detection advantages were most consistent relative to WMH-SynthSeg; SHIVA-WMH and segcsvdWMH detected small lesions at similar rates in this 3D set, although SHIVA-WMH generated more very small false-positive lesions (5.04 vs. 0.84 per scan). Representative segmentations across the range of WMH burden are shown in **Figure 3C**.

**Figure 3.**
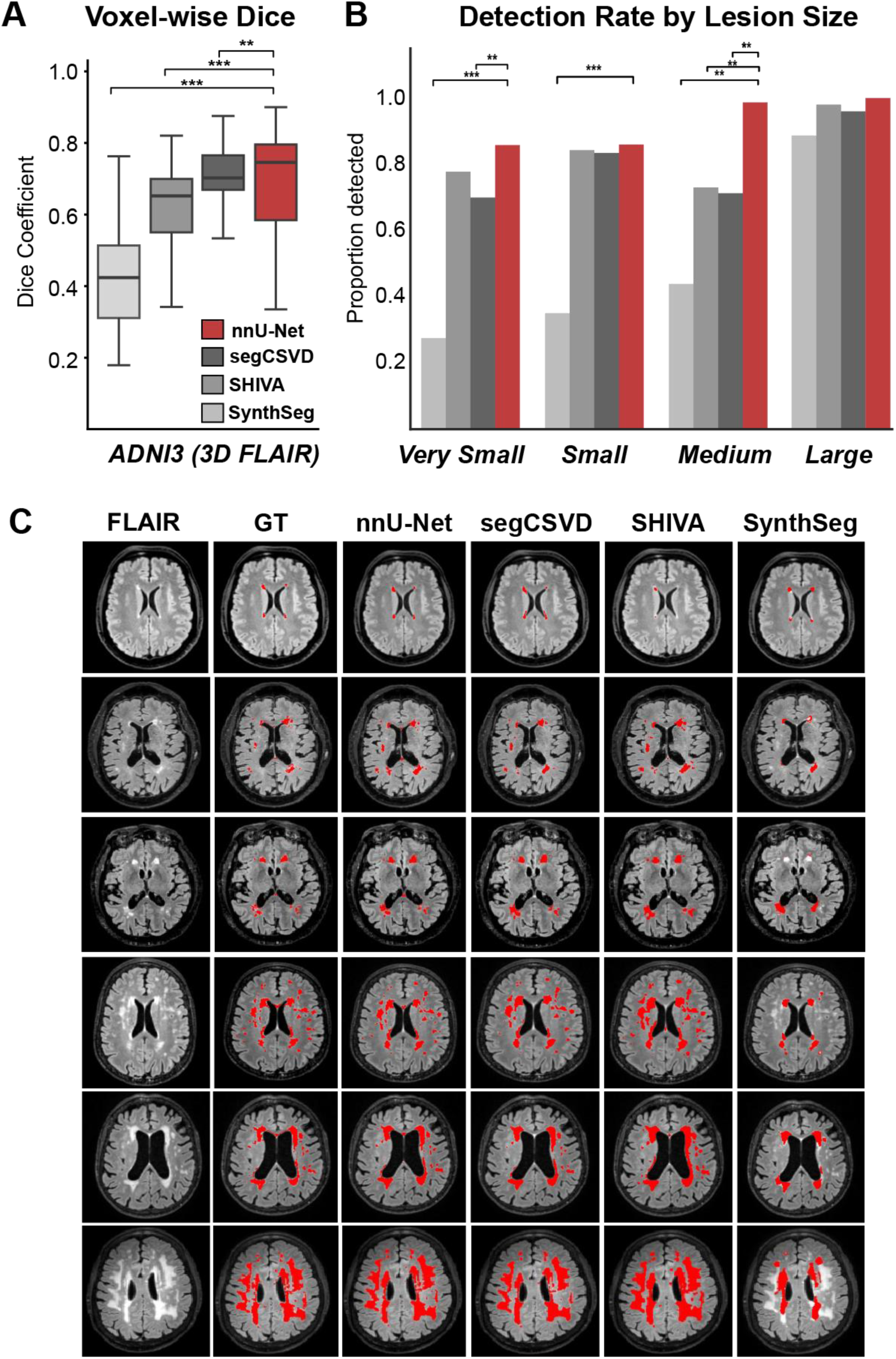
External validation in an independent ADNI3 3D FLAIR set (n = 25). **A.** Participant-level voxel-wise Dice for each method (n = 25). **B.** Lesion detection rate by reference lesion volume category. **C.** Representative axial FLAIR images, reference masks (GT), and predicted masks (red overlay) from each method in six participants ordered by increasing reference WMH burden (top to bottom). The ADNI3 set was not used for training, model selection, or threshold tuning. Box-plot conventions, color scheme, and statistical testing are as in Figure 2; \*\**p* < 0.01, \*\*\**p* < 0.001. Abbreviations: ADNI, Alzheimer’s Disease Neuroimaging Initiative; FLAIR, fluid-attenuated inversion recovery; GT, ground truth (reference mask); WMH, white matter hyperintensity.

### WMH volume and incident dementia

Among 1,174 cognitively unimpaired participants from ADNI and the Knight ADRC, 42 progressed to dementia over a median follow-up of 5.9 years (**Table 3**). After adjustment for age, sex, education, *APOE* ε4 carrier status, and cohort, higher WMH volume from the proposed model was associated with incident dementia (OR per SD of WMH volume 1.40, 95% CI 1.11– 1.77) and improved model fit over the clinical model (likelihood-ratio χ^2^ = 6.90, *p* = 0.009; **Table 3**; **Figure 4**). Cross-validated AUC increased from 0.683 for the clinical model to 0.712 (95% CI 0.62–0.79; ΔAUC = 0.029, permutation *p* < 0.001). WMH volumes from WMH-SynthSeg (OR 1.30, 95% CI 1.02–1.67) and SHIVA-WMH (OR 1.27, 95% CI 1.01–1.59) showed weaker associations, and neither significantly improved model fit for incidence dementia beyond clinical covariates (likelihood-ratio *p* = 0.052 and 0.059; ΔAUC = 0.015 and 0.011). segcsvdWMH-derived volume was not associated with incident dementia (OR 1.07, 95% CI 0.81–1.41; likelihood-ratio *p* = 0.65; ΔAUC = −0.005). After additional adjustment for FLAIR acquisition type, WMH volume from the proposed model remained associated with incident dementia (OR per SD 1.34, 95% CI 1.05–1.70; likelihood-ratio *p* = 0.027; **Supplemental Table S6**).

**Figure 4.**
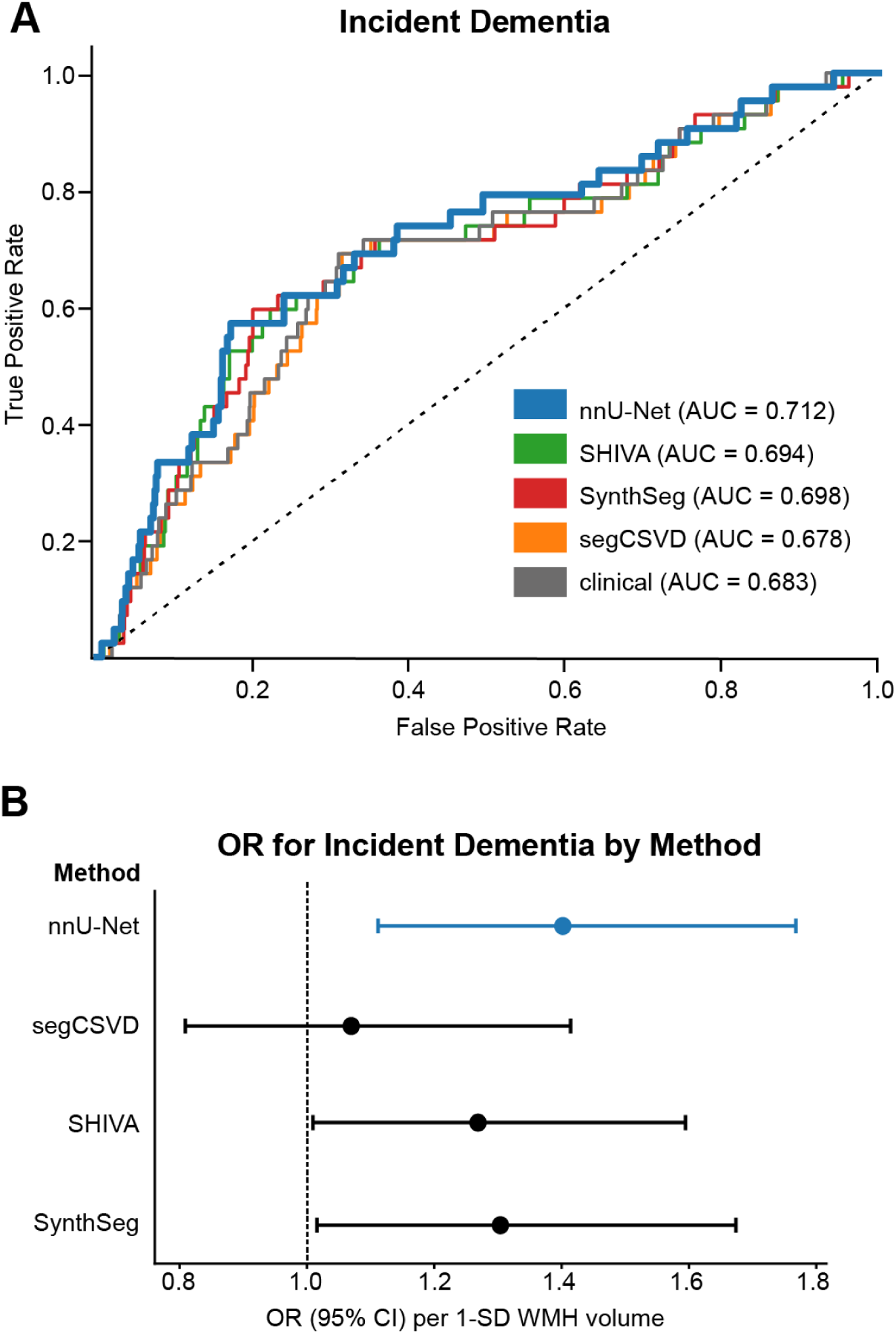
Algorithm-derived WMH volumes and incident dementia. Analyses included 1,174 cognitively unimpaired ADNI and Knight ADRC participants (global CDR = 0), including n = 42 who progressed to CDR ≥1. **A.** Ten-fold cross-validated ROC curves for the clinical model (age, sex, education, *APOE ε*4 status, and cohort) and models adding standardized WMH volume from each segmentation method, using identical folds. **B.** Adjusted OR (95% CI) for incident dementia per 1-SD increase in WMH volume; dashed line indicates OR = 1. Abbreviations: AUC, area under the curve; CI, confidence interval; OR, odds ratio; WMH, white matter hyperintensity.

**Table 3.**
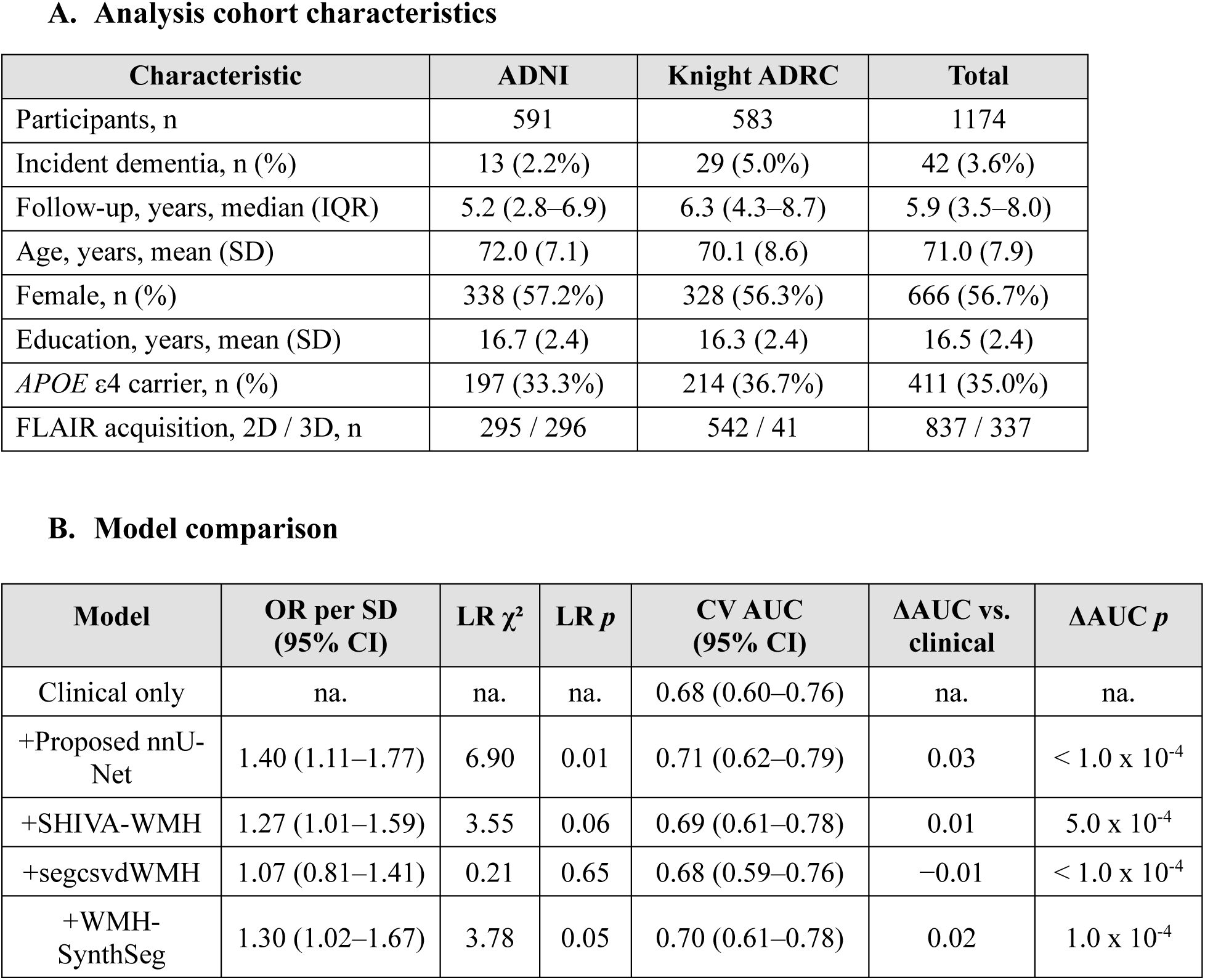
Participant characteristics and model comparisons for incident dementia. **A.** Baseline characteristics of cognitively unimpaired participants (global CDR = 0). **B.** Logistic regression models for incident dementia (global CDR ≥ 1 during follow-up). The clinical model included age, sex, years of education, *APOE* ε4 carrier status, and cohort; each WMH model additionally included standardized WMH volume from one segmentation method. ORs are per 1-SD increase in WMH volume. LR tests and ΔAUC compared WMH models with the clinical-only model. AUCs were estimated using identical 10-fold cross-validation splits, with bootstrap 95% CIs and permutation *p* values for ΔAUC. AUC, area under the receiver operating characteristic curve; CDR, Clinical Dementia Rating; CI, confidence interval; LR, likelihood ratio; OR, odds ratio.

| **A. Analysis cohort characteristics** |  |  |  |  |  |  |
| **Characteristic** | **ADNI** | **Knight ADRC** | **Total** |  |  |  |
|---|---|---|---|---|---|---|
| Participants, n | 591 | 583 | 1174 |  |  |  |
| Incident dementia, n (%) | 13 (2.2%) | 29 (5.0%) | 42 (3.6%) |  |  |  |
| Follow-up, years, median (IQR) | 5.2 (2.8–6.9) | 6.3 (4.3–8.7) | 5.9 (3.5–8.0) |  |  |  |
| Age, years, mean (SD) | 72.0 (7.1) | 70.1 (8.6) | 71.0 (7.9) |  |  |  |
| Female, n (%) | 338 (57.2%) | 328 (56.3%) | 666 (56.7%) |  |  |  |
| Education, years, mean (SD) | 16.7 (2.4) | 16.3 (2.4) | 16.5 (2.4) |  |  |  |
| *APOE* ε4 carrier, n (%) | 197 (33.3%) | 214 (36.7%) | 411 (35.0%) |  |  |  |
| FLAIR acquisition, 2D / 3D, n | 295 / 296 | 542 / 41 | 837 / 337 |  |  |  |
| **B. Model comparison** |  |  |  |  |  |  |
| **Model** | **OR per SD<br>(95% CI)** | **LR χ²** | **LR *p*** | **CV AUC<br>(95% CI)** | **ΔAUC vs. clinical** | **ΔAUC *p*** |
| Clinical only | na. | na. | na. | 0.68 (0.60–0.76) | na. | na. |
| +Proposed nnU-Net | 1.40 (1.11–1.77) | 6.90 | 0.01 | 0.71 (0.62–0.79) | 0.03 | < 1.0 x 10<sup>-4</sup> |
| +SHIVA-WMH | 1.27 (1.01–1.59) | 3.55 | 0.06 | 0.69 (0.61–0.78) | 0.01 | 5.0 x 10<sup>-4</sup> |
| +segcsvdWMH | 1.07 (0.81–1.41) | 0.21 | 0.65 | 0.68 (0.59–0.76) | −0.01 | < 1.0 x 10<sup>-4</sup> |
| +WMH-SynthSeg | 1.30 (1.02–1.67) | 3.78 | 0.05 | 0.70 (0.61–0.78) | 0.02 | 1.0 x 10<sup>-4</sup> |

## DISCUSSION

We developed a single nnU-Net WMH segmentation model trained across neurodegenerative, CSVD-enriched, and population-based cohorts and across conventional 2D and isotropic 3D FLAIR. Four findings support its use as a common segmentation approach in multi-cohort studies. First, the model outperformed three contemporary tools at the voxel, volume, and lesion levels in a held-out internal test set. Second, accuracy was similar on 2D and 3D FLAIR, and volume bias relative to acquisition-specific references varied less between paired acquisitions than comparator biases. Third, its advantage was largest where WMH measurement is most difficult: in low-burden scans and for small lesions. Fourth, performance transferred to an unseen ADNI3 3D FLAIR protocol, and model-derived volumes added information about incident dementia beyond clinical covariates. The principal contribution is therefore not the nnU-Net architecture itself, but the development and validation of a single model across heterogeneous disease contexts and acquisitions, together with evidence that algorithm-dependent differences in WMH quantification can affect downstream clinical inference.

Acquisition-related segmentation bias is an important consideration in multi-cohort studies. Differences in error between 2D and 3D FLAIR can complicate comparisons when acquisition type varies by cohort, site, or study phase. The same-visit paired WWISH scans allowed us to assess these differences within participants. The proposed model’s mean volume bias relative to acquisition-specific references differed by less than 2 mL between acquisitions, compared with approximately 5–10 mL for the comparators; SHIVA-WMH also showed reversals in bias direction. These findings are consistent with Hotz et al. (2022), who reported different algorithm biases on 2D and 3D FLAIR relative to acquisition-specific manual references. Expert reference volumes themselves were, on average, 6.3 mL larger on 2D than on 3D FLAIR, and the proposed model’s volumes differed in the same direction. Thus, similar accuracy against acquisition-specific references does not establish interchangeability of 2D- and 3D-derived volumes.

The largest gains were in low-burden scans and small lesions, where existing tools differed most from the reference. This range matters because many population-based and preclinical samples, such as UKB, have total WMH volumes below 5 mL, where measurement error is large relative to between-person variation. The proposed model’s higher sensitivity did not come at the cost of more false-positive lesions than SHIVA-WMH, and the low false-positive count of segcsvdWMH reflected conservative segmentation that missed many small lesions. SHIVA-WMH, which was developed with isotropic 3D FLAIR to detect punctate lesions (Tsuchida et al., 2024), matched the proposed model’s very-small-lesion overlap on 3D FLAIR but not on 2D FLAIR, consistent with the importance of training-data acquisition for small-lesion detection. More reliable detection at low burden may increase the usefulness of WMH as an early marker in cognitively unimpaired and population samples, although this requires direct testing in longitudinal data.

These comparisons should be interpreted in light of each comparator’s design. segcsvdWMH was trained on heterogeneous 2D and 3D FLAIR from CSVD-related clinical cohorts (Gibson et al., 2024); SHIVA-WMH was trained on isotropic 3D FLAIR from young adults combined with the MICCAI WMH Segmentation Challenge data (Tsuchida et al., 2024); and WMH-SynthSeg was designed to be contrast- and resolution-agnostic across clinical and low-field MRI rather than to maximize accuracy on research-grade FLAIR (Laso et al., 2024). Input requirements also differed: our proposed model and SHIVA-WMH used both FLAIR and T1-weighted images, whereas segcsvdWMH and WMH-SynthSeg were run using FLAIR images alone. Comparator Dice values in our data were lower than those reported in their development studies. This likely reflects, in part, differences between our reference labeling convention and theirs, in addition to differences in populations and protocols. Because the proposed model was trained on masks generated with the same convention used for evaluation, voxel-level benchmarking alone may favor it. The incident dementia analysis does not depend on reference masks and therefore provides a complementary, label-independent comparison. The proposed model also exceeded segcsvdWMH within WWISH on both 2D and 3D FLAIR, although the margin was smaller on 3D (0.74 vs. 0.66), and segcsvdWMH was the closest competitor in ADNI3, with similar volumetric error to the proposed model.

Algorithm choice also influenced downstream clinical inference. All four methods were applied to the same participants and evaluated using the same clinical covariates, yet only WMH volume from the proposed model significantly improved model fit for incident dementia. Volumes from SHIVA-WMH and WMH-SynthSeg showed weaker associations, and segcsvdWMH-derived volume showed no association. These results extend prior evidence that WMH–cognition associations depend on the segmentation algorithm (Tubi et al., 2020). However, absolute AUC gains were modest. These findings provide complementary evidence of the clinical relevance of the proposed model’s WMH estimates; they do not establish a validated clinical prediction tool. Its stronger performance in low-burden scans and small lesions may contribute to the observed associations, although this explanation requires direct testing. The dementia analysis combined 2D and 3D FLAIR scans from ADNI and Knight ADRC; additional adjustment for acquisition type did not change the association for the proposed model.

For multi-cohort studies, these results support applying a single segmentation model across the evaluated disease contexts and acquisitions, reducing the need to switch algorithms between cohorts. This removes algorithm choice as one source of methodological variation, but does not eliminate acquisition-related differences in WMH volume. Acquisition type should therefore remain part of the analytical design, and comparable segmentation accuracy should not be taken to imply that unadjusted 2D- and 3D-derived volumes can be pooled interchangeably. Future work could evaluate whether incorporating paired acquisitions during training improves cross-acquisition volume consistency while preserving segmentation accuracy.

This study has several strengths, including deliberately heterogeneous training data, same-visit paired 2D and 3D acquisitions, head-to-head benchmarking at the voxel, volume, and lesion levels, evaluation in an unseen acquisition protocol, and a label-independent clinical validation. Several limitations warrant consideration. First, although the cohorts were diverse, they do not capture the full range of scanners, field strengths, acquisition protocols, and populations encountered in practice. Second, reference masks were initialized using an automated method and refined by a single rater with neurologist adjudication. Inter-rater Dice agreement, previously assessed in a subset, was 0.72 (95% CI, 0.67–0.81; Phuah et al., 2022), comparable to the model’s mean Dice in the internal test set (0.73), suggesting that model–reference agreement approached the level of agreement between human raters. However, reliability was not reassessed here, and systematic labeling conventions may be shared by training and evaluation. Third, the external ADNI3 test set included distinct participants and acquisition protocol but originated from the same consortium as part of the development data, and its size was modest (n = 25), although comparable to or larger than the labeled external test sets used to validate SHIVA-WMH (n = 11) and segcsvdWMH (n = 10); fully independent validation in larger cohorts remains necessary. Fourth, paired WWISH acquisitions contributed correlated observations to pooled analyses, although results were consistent in acquisition-specific strata in which each participant contributed one scan. Fifth, reference masks were defined separately for each acquisition, allowing assessment of segmentation accuracy but not determination of which acquisition more accurately measured the underlying lesion burden. Direct cross-acquisition agreement was also assessed in only 26 participants from a single cohort. Sixth, the model requires both FLAIR and a coregistered T1-weighted image, which may limit its use in datasets without T1-weighted imaging or with poor FLAIR–T1 registration. It was also evaluated cross-sectionally; its accuracy for measuring longitudinal change and its regional performance remain to be established. Finally, clinical validation was limited to progression on the global CDR in a modest sample.

Pooled and longitudinal WMH studies combine cohorts that differ in disease composition and FLAIR protocol, creating the potential for acquisition-related measurement differences to be mistaken for biological differences between cohorts, sites, or study phases. Our findings show that algorithm choice matters: segmentation methods differed in acquisition-dependent bias and in the clinical information their WMH estimates provided. A single model trained across disease contexts and both 2D and 3D FLAIR achieved similar segmentation accuracy on both acquisitions and more stable volume bias relative to acquisition-specific references than the comparator tools. These results support using a common segmentation approach across heterogeneous cohorts, reducing the need to exclude scans or switch algorithms solely because of acquisition type. Public release of the model will facilitate its use in multi-cohort aging, vascular, and neurodegenerative studies, including studies that span protocol transitions.

### Ethics Statement

The present analysis was determined to be exempt from institutional review board review by the Dignity Health Arizona Research Enterprise (DHARE). The parent studies were approved by their respective institutional review boards, and written informed consent was obtained from participants or their legally authorized representatives, as applicable.

## Data Availability

ADNI and UK Biobank data are available through their respective data-access procedures. Access to WWISH and Knight ADRC data is subject to institutional approval and applicable data-use agreements.

## Funding

This work was supported by grants from the National Institute of Health (NIH): R01AG089239 (CP), RF1NS116565 (AF) and R01HL129241 (AF). The Knight-ADRC imaging resources used in this study were supported by the NIH under award number P30NS048056. Data collection and sharing for ADNI is funded by the NIH Health Grant U19 AG024904. The funders had no role in study design, data analysis, interpretation or writing of the manuscript.

## Authors’ contributions

YC and UK contributed equally. YC developed the WMH segmentation pipeline. YC and UK processed imaging data. UK performed statistical analyses and contributed to interpretation and manuscript drafting. BX curated imaging and phenotype data and performed WMH quality control. BX and CP created the manual reference masks. CP conceived and designed the study, supervised analyses, interpreted results, and drafted the manuscript. HA and AF contributed WWISH imaging data, and SA curated WWISH clinical data. JML contributed to study design and interpretation. All authors critically revised and approved the final manuscript.

## Conflicts of Interest

The authors have nothing to report.

## Data and Code Availability Statement

ADNI and UK Biobank data are available through their respective data-access procedures. Access to WWISH and Knight ADRC data is subject to institutional approval and applicable data-use agreements. The trained model and containerized preprocessing and segmentation pipeline are publicly available at https://github.com/uttamkhatri/anyflair.

## Acknowledgements

Data used in the preparation of this article were obtained from the Alzheimer’s Disease Neuroimaging Initiative (ADNI) database. ADNI investigators contributed to the design and implementation of ADNI and/or provided data but did not participate in the analysis or writing of this report. This research has been conducted using the UK Biobank Resource under Application Number 122019.

## Notes

### Competing Interest Statement

The authors have declared no competing interest.

## REFERENCES

Balakrishnan, R., Valdés Hernández, M. del C., & Farrall, A. J. (2021). Automatic segmentation of white matter hyperintensities from brain magnetic resonance images in the era of deep learning and big data – A systematic review. Computerized Medical Imaging and Graphics, 88, 101867. 10.1016/j.compmedimag.2021.101867

Caligiuri, M. E., Perrotta, P., Augimeri, A., Rocca, F., Quattrone, A., & Cherubini, A. (2015). Automatic Detection of White Matter Hyperintensities in Healthy Aging and Pathology Using Magnetic Resonance Imaging: A Review. Neuroinformatics, 13(3), 261–276. 10.1007/s12021-015-9260-y

DeCarli, C., Rajan, K. B., Jin, L.-W., Hinman, J., Johnson, D. K., Harvey, D., Fornage, M., & on behalf of the Diverse VCID Study Investigators. (2025). WMH Contributions to Cognitive Impairment: Rationale and Design of the Diverse VCID Study. Stroke, 56(3), 758–776. 10.1161/STROKEAHA.124.045903

Ganiler, O., Oliver, A., Diez, Y., Freixenet, J., Vilanova, J.C., Beltran, B., Ramió-Torrentà, L., Rovira, A., Lladó, X. (2014). A subtraction pipeline for automatic detection of new appearing multiple sclerosis lesions in longitudinal studies. Neuroradiology, 56(5), 363–74. doi: 10.1007/s00234-014-1343-1.

Garnier-Crussard, A., Cotton, F., Krolak-Salmon, P., & Chételat, G. (2023). White matter hyperintensities in Alzheimer’s disease: Beyond vascular contribution. Alzheimer’s & Dementia, 19(8), 3738–3748. 10.1002/alz.13057

Gesierich, B., Pirpamer, L., Meier, D. S., Amann, M., Cerfontaine, M. N., de Leeuw, F.-E., Maillard, P., Moy, S., Helmer, K. G., Kühne, M., Bonati, L. H., Rutten, J. W., Lesnik Oberstein, S. A. J., & Duering, M. (2025). Technical and clinical validation of a novel deep learning-based white matter hyperintensity segmentation tool. Cerebral Circulation-Cognition and Behavior, 9, 100393. 10.1016/j.cccb.2025.100393

Gibson, E., Ramirez, J., Woods, L. A., Ottoy, J., Berberian, S., Scott, C. J. M., Yhap, V., Gao, F., Coello, R. D., Valdes Hernandez, M., Lang, A. E., Tartaglia, C. M., Kumar, S., Binns, M. A., Bartha, R., Symons, S., Swartz, R. H., Masellis, M., Singh, N., … ONDRI Investigators, ADNI, CAIN Investigators, colleagues from the Foundation Leducq Transatlantic Network of Excellence. (2024). segcsvdWMH: A Convolutional Neural Network-Based Tool for Quantifying White Matter Hyperintensities in Heterogeneous Patient Cohorts. Human Brain Mapping, 45(18), e70104. 10.1002/hbm.70104

Heinen, R., Steenwijk, M. D., Barkhof, F., Biesbroek, J. M., van der Flier, W. M., Kuijf, H. J., Prins, N. D., Vrenken, H., Biessels, G. J., & de Bresser, J. (2019). Performance of five automated white matter hyperintensity segmentation methods in a multicenter dataset. Scientific Reports, 9(1), 16742. 10.1038/s41598-019-52966-0

Hoopes, A., Mora, J.S., Dalca, A.V., Fischl, B., Hoffmann, M. (2022). SynthStrip: skull-stripping for any brain image. Neuroimage, 10(15),260:119474. doi: 10.1016/j.neuroimage.2022.119474.

Hotz, I., Deschwanden, P. F., Liem, F., Mérillat, S., Malagurski, B., Kollias, S., & Jäncke, L. (2022). Performance of three freely available methods for extracting white matter hyperintensities: FreeSurfer, UBO Detector, and BIANCA. Human Brain Mapping, 43(5), 1481–1500. 10.1002/hbm.25739

Isensee, F., Jaeger, P. F., Kohl, S. A. A., Petersen, J., & Maier-Hein, K. H. (2021). nnU-Net: A self-configuring method for deep learning-based biomedical image segmentation. Nature Methods, 18(2), 203–211. 10.1038/s41592-020-01008-z

Jenkinson, M., Bannister, P., Brady, M., Smith, S. (2002). Improved optimization for the robust and accurate linear registration and motion correction of brain images. NeuroImage,17(2),825–841. doi: 10.1006/nimg.2002.1132

Kuijf, H. J., Biesbroek, J. M., De Bresser, J., Heinen, R., Andermatt, S., Bento, M., Berseth, M., Belyaev, M., Cardoso, M. J., Casamitjana, A., Collins, D. L., Dadar, M., Georgiou, A., Ghafoorian, M., Jin, D., Khademi, A., Knight, J., Li, H., Lladó, X., … Biessels, G. J. (2019). Standardized Assessment of Automatic Segmentation of White Matter Hyperintensities and Results of the WMH Segmentation Challenge. IEEE Transactions on Medical Imaging, 38(11), 2556–2568. 10.1109/TMI.2019.2905770

Laso, P., Cerri, S., Sorby-Adams, A., Guo, J., Mateen, F., Goebl, P., Wu, J., Liu, P., Li, H., Young, S. I., Billot, B., Puonti, O., Sze, G., Payabavash, S., DeHavenon, A., Sheth, K. N., Rosen, M. S., Kirsch, J., Strisciuglio, N., … Iglesias, J. E. (2024). Quantifying white matter hyperintensity and brain volumes in heterogeneous clinical and low-field portable MRI. Proc IEEE Int Symp Biomed Imaging, 10.1109/isbi56570.2024.10635502. doi: 10.1109/isbi56570.2024.10635502.

Maillard, P., Fletcher, E., Carmichael, O., Schwarz, C., Seiler, S., DeCarli, C., & Alzheimer’s Disease Neuroimaging Initiative. (2024). Cerebrovascular markers of WMH and infarcts in ADNI: A historical perspective and future directions. Alzheimer’s & Dementia: The Journal of the Alzheimer’s Association, 20(12), 8953–8968. 10.1002/alz.14358

Morris, J. C., Ernesto, C., Schafer, K., Coats, M., Leon, S., Sano, M., Thal, L. J., & Woodbury, P. (1997). Clinical Dementia Rating training and reliability in multicenter studies: The Alzheimer’s Disease Cooperative Study experience. Neurology, 48(6), 1508–1510. 10.1212/WNL.48.6.1508

Phuah, C.-L., Chen, Y., Strain, J. F., Yechoor, N., Laurido-Soto, O. J., Ances, B. M., Lee, J.-M., & Alzheimer’s Disease Neuroimaging Initiative. (2022). Association of data-driven white matter hyperintensity spatial signatures with distinct cerebral small vessel disease etiologies. Neurology, 99(23), e2535–e2547. 10.1212/WNL.0000000000201186

Rahmani, M., Dierker, D., Yaeger, L., Saykin, A., Luckett, P. H., Vlassenko, A. G., Owens, C., Jafri, H., Womack, K., Fripp, J., Xia, Y., Tosun, D., Benzinger, T. L. S., Masters, C. L., Lee, J.-M., Morris, J. C., Goyal, M. S., Strain, J. F., Kukull, W., … Artificial Intelligence and Machine Learning. (2024). Evolution of white matter hyperintensity segmentation methods and implementation over the past two decades; an incomplete shift towards deep learning. Brain Imaging and Behavior, 18(5), 1310–1322. 10.1007/s11682-024-00902-w

Smith, E. E., & Beaudin, A. E. (2018). New insights into cerebral small vessel disease and vascular cognitive impairment from MRI. Current Opinion in Neurology, 31(1), 36–43. 10.1097/WCO.0000000000000513

Torres-Simon, L., del Cerro-León, A., Yus, M., Bruña, R., Gil-Martinez, L., Dolado, A. M., Maestú, F., Arrazola-Garcia, J., & Cuesta, P. (2024). Decoding the best automated segmentation tools for vascular white matter hyperintensities in the aging brain: A clinician’s guide to precision and purpose. GeroScience, 46(6), 5485–5504. 10.1007/s11357-024-01238-5

Tsuchida, A., Boutinaud, P., Verrecchia, V., Tzourio, C., Debette, S., & Joliot, M. (2024). Early detection of white matter hyperintensities using SHIVA-WMH detector. Human Brain Mapping, 45(1), e26548. 10.1002/hbm.26548

Tubi, M. A., Feingold, F. W., Kothapalli, D., Hare, E. T., King, K. S., Thompson, P. M., & Braskie, M. N. (2020). White matter hyperintensities and their relationship to cognition: Effects of segmentation algorithm. NeuroImage, 206, 116327. 10.1016/j.neuroimage.2019.116327

Tustison, N.J., Avants, B.B., Cook, P.A., Zheng, Y., Egan, A., Yushkevich, P.A., Gee, J.C. (2010) N4ITK: Improved N3 bias correction. IEEE Transactions on Medical Imaging, 29(6),1310–1320. doi: 10.1109/TMI.2010.2046908

Vemuri, P., Decarli, C., & Duering, M. (2022). Imaging Markers of Vascular Brain Health: Quantification, Clinical Implications, and Future Directions. Stroke, 53(2), 416–426. 10.1161/STROKEAHA.120.032611

Wardlaw, J. M., Valdés Hernández, M. C., & Muñoz-Maniega, S. (2015). What are White Matter Hyperintensities Made of? Journal of the American Heart Association: Cardiovascular and Cerebrovascular Disease, 4(6), e001140. 10.1161/JAHA.114.001140

