## Supplementary Method & Figures for "A Multi-Cohort nnU-Net for White Matter Hyperintensity Segmentation Across 2D and 3D FLAIR Acquisitions"

**SUPPLEMENTARY METHODS - Cohort descriptions**

**ADNI.** The Alzheimer's Disease Neuroimaging Initiative (ADNI) is an ongoing, longitudinal, multicenter observational study designed to develop and validate clinical, imaging, genetic, and biochemical biomarkers for the early detection and tracking of Alzheimer's disease (AD). Across multiple phases (ADNI1, GO, 2, 3 and 4), the study enrolled adults aged approximately 55-90 years into cognitively unimpaired, mild cognitive impairment (MCI), and dementia/Alzheimer's disease diagnostic cohorts, using standardized clinical criteria and neuropsychological assessments to define group membership.[1] Participants undergo serial structural MRI, PET, CSF, and blood biomarker collection alongside longitudinal cognitive and functional testing, enabling characterization of biomarker trajectories from normal aging through prodromal and clinical AD and providing a harmonized multi-site imaging resource for AD-spectrum pathology. In this study, ADNI contributes T1-weighted and FLAIR MRI with varying degrees of neurodegenerative and vascular lesion burden across the AD diagnostic spectrum, allowing evaluation of segmentation performance in mixed neurodegenerative-vascular pathology.

**WWISH.** The WashU White Matter ISchemic Hypoxia (WWISH) cohort is a single-center study established at Washington University in St. Louis to characterize cerebral small vessel disease (CSVD) in patients with clinically defined CSVD and elevated vascular risk factor burden (e.g. hypertension, diabetes, hyperlipidemia). It included individuals referred for neuroimaging, with CSVD diagnosis supported by clinical criteria and MRI evidence of white matter hyperintensities (WMH), lacunes, and other CSVD markers, and is designed to capture a spectrum of lesion burden from mild to severe. The imaging protocol acquires paired 2D FLAIR, 3D FLAIR, and T1-weighted MRI at the same visit (n=130), providing cross-sequence data at identical timepoints to directly assess protocol-dependent variability in WMH measurement.

**UKB.** The UK Biobank (UKB) is a large, prospective, population-based cohort of ~500,000 volunteers aged 40-69 years recruited across the United Kingdom between 2006 and 2010, with extensive baseline phenotyping and long-term follow-up through linked electronic health records and death registries. A major imaging enhancement is acquiring multimodal MRI in a subset of participants (target n=100,000), including structural T1-weighted, 3D FLAIR, diffusion, and functional brain MRI, together with cardiac, abdominal, musculoskeletal, and vascular imaging.[2] These imaging data, combined with detailed vascular risk profiles, cognitive assessments, and incident disease outcomes, provide a population-level resource for studying the distribution, determinants, and clinical consequences of WMH and other markers of CSVD in predominantly cognitively normal middle-aged and older adults. In this study, we leverage the UKB brain MRI subset with available T1-weighted and 3D FLAIR sequences (n=100) to train and evaluate WMH segmentation models in a population-based setting with heterogeneous vascular risk exposure and largely subclinical lesion burden.

**Knight ADRC.** The Charles F. and Joanne Knight Alzheimer Disease Research Center (Knight ADRC) at Washington University in St. Louis conducts longitudinal studies of aging and Alzheimer disease, enrolling community-dwelling adults across the spectrum from normal cognition to symptomatic disease. Participants undergo repeated clinical and cognitive assessments, with neuroimaging, genetic, and fluid biomarker characterization available through affiliated research programs. Clinical evaluations incorporate medical and neurological assessment and interviews with participants and knowledgeable collateral sources to characterize changes in cognition and everyday functioning. Assessments follow standardized protocols, including the National Alzheimer's Coordinating Center Uniform Data Set, and cognitive and functional status is staged using the Clinical Dementia Rating (CDR). For the present study, we included participants who were cognitively unimpaired at baseline (global CDR = 0), had an eligible FLAIR scan, and underwent subsequent clinical follow-up. Incident dementia was defined as progression to a global CDR ≥ 1 during follow-up.

### References


1. Aisen PS, Petersen RC, Donohue MC, Gamst A, Raman R, Thomas RG, et al. Clinical Core of the Alzheimer's Disease Neuroimaging Initiative: progress and plans. *Alzheimers Dement.* 2010 May;6(3):239-46.

2. Sudlow C, Gallacher J, Allen N, Beral V, Burton P, Danesh J, et al. UK biobank: an open access resource for identifying the causes of a wide range of complex diseases of middle and old age. *PLoS Med*. 2015 Mar 31;12(3):e1001779.

**Supplementary Table S1.** FLAIR and T1-weighted acquisition parameters by cohort. All scans were acquired at 3T; ranges reflect variation across sites or scanners. ADNI 2D FLAIR was used for model development and ADNI3 3D FLAIR for external validation. Abbreviations: TE, echo time; TI, inversion time; TR, repetition time; ADNI, Alzheimer's Disease Neuroimaging Initiative; FLAIR, fluid-attenuated inversion recovery; MPRAGE, magnetization-prepared rapid acquisition gradient echo; IR-FSPGR, inversion-recovery fast spoiled gradient echo; UKB, UK Biobank; WWISH, Washington University White Matter Ischemic-Hypoxia study.

| Cohort | FLAIR Type | TE (ms) | TR (ms) | TI (ms) | Flip Angle (°) | Resolution (mm) | Slice Thickness (mm) |
|---|---|---|---|---|---|---|---|
| ADNI | 2D | 90—91 | 9000 | 2500 | 90—150 | 0.83 × 0.83 to 0.88 × 0.88 | 5.0 |
| ADNI | 3D | 116—442 | 4800—4892 | 0—1650 | 90—120 | 1.00 × 1.00 | 1.2 |
| WWISH | 2D | 94 | 9000 | 2500 | 150 | 0.86 × 0.86 | 3.0 |
| WWISH | 3D | 417 | 4800 | 1650 | — | 1.00 × 1.00 | 1.0 |
| UKB | 3D | 395 | 5000 | 1800 | — | 1.00 × 1.00 | 1.05 |

| Cohort | T1-weighted sequence | TE (ms) | TR (ms) | TI (ms) | Flip Angle (°) | Resolution (mm) | Slice Thickness (mm) |
|---|---|---|---|---|---|---|---|
| ADNI | MPRAGE / IR-FSPGR | 3.0 / 2.8—3.2 | 2300 / 7—8 | 900 / 400 | — | 1.0 × 1.0 × 1.2 | 1.2 |
| ADNI (external test) | MPRAGE / IR-FSPGR | 3.0 / 2.9—3.2 | 2300 / 6—8 | 900 / 400 | — | 1.0 × 1.0 × 1.0 | 1.0 |
| WWISH | 3D MPRAGE | 2.62 | 2400 | 1000 | 8 | 1.0 × 1.0 × 1.0 | 1.0 |
| UKB | 3D MPRAGE | 2.01 | 2000 | 880 | 8 | 1.0 × 1.0 × 1.0 | 1.0 |

**Supplementary Figure S1. Bland-Altman agreement between predicted and reference total WMH volume.** The x-axis shows the mean of predicted and reference volume (mL); the y-axis shows the difference (predicted − reference, mL). **A.** Internal held-out test set (92 scans). **B.** External ADNI3 3D FLAIR set (n = 25). **C.** Internal test set by FLAIR acquisition (2D, n = 46; 3D, n = 46). **D.** Internal test set by cohort and acquisition (ADNI 2D, n = 20; UKB 3D, n = 20; WWISH 2D, n = 26; WWISH 3D, n = 26). Red diamonds, proposed nnU-Net model; dark gray squares, segcsvdWMH; gray triangles, SHIVA-WMH; light gray circles, WMH-SynthSeg. Solid horizontal lines indicate mean bias and dotted lines the 95% limits of agreement (mean ± 1.96 SD) for each method; red lines correspond to the proposed model. GT, ground truth (reference mask); WMH, white matter hyperintensity.

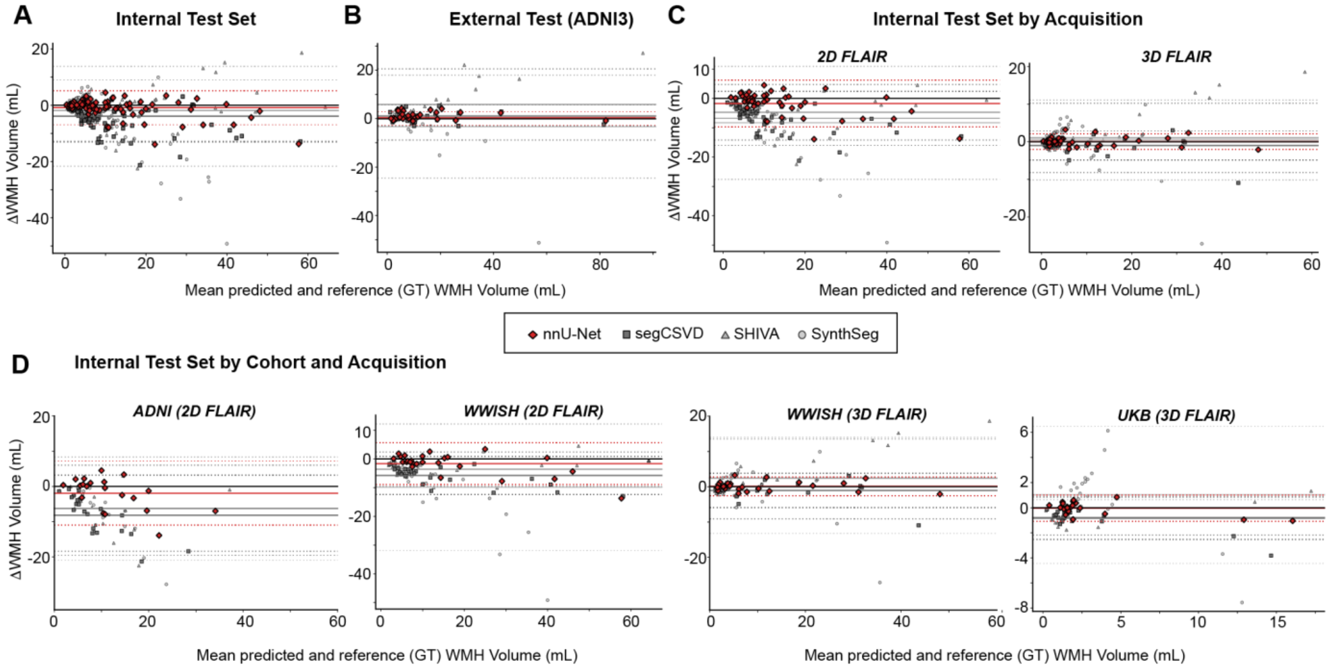

**Supplementary Figure S2. Lesion-level performance by cohort and FLAIR acquisition in the internal test set. A.** Lesion-wise Dice (left) and lesion detection rate (right) by reference lesion volume category for ADNI 2D (n = 20), UKB 3D (n = 20), WWISH 2D (n = 26), and WWISH 3D (n = 26). **B.** The same metrics by FLAIR acquisition (2D, n = 46; 3D, n = 46).

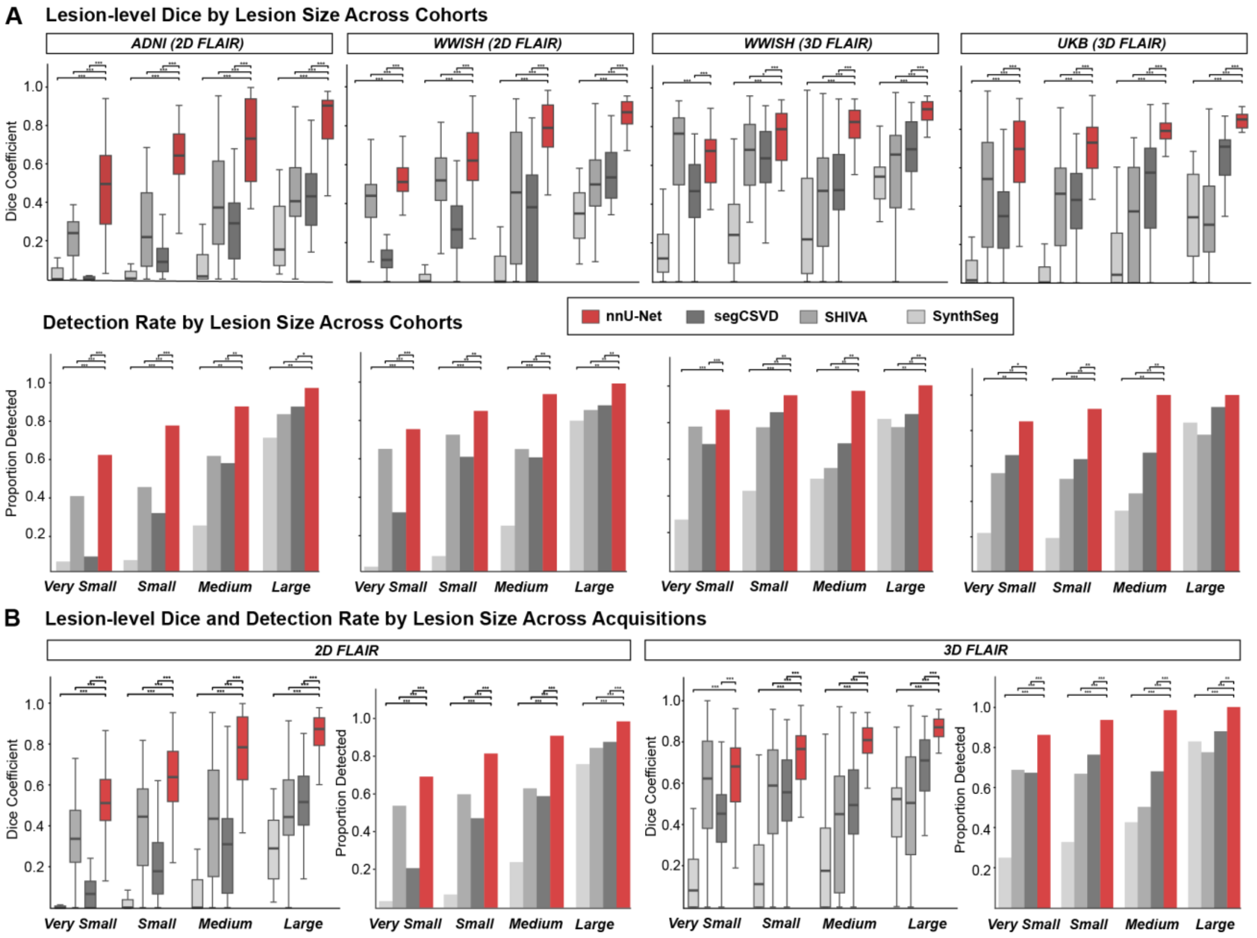
